# Whole-exome sequencing analysis on products of conception: A cohort study to evaluate clinical utility and genetic etiology for pregnancy loss

**DOI:** 10.1101/2020.07.19.20150144

**Authors:** Chen Zhao, Hongyan Chai, Qinghua Zhou, Jiadi Wen, Uma M. Reddy, Rama Kastury, Yonghui Jiang, Winifred Mak, Allen E. Bale, Hui Zhang, Peining Li

**Affiliations:** Departments of Genetics, Yale School of Medicine, New Haven, CT, USA; Obstetrics and Gynecology, Yale School of Medicine, New Haven, CT, USA; Department of Women’s Health, Dell Medical School, The University of Texas at Austin, TX, USA; Biomedical Translational Research Institute, Jinan University, Guangzhou, China

**Keywords:** whole exome sequencing (WES), pregnancy loss, products of conception (POC), abnormality detection rate (ADR), genetic etiology

## Abstract

**Purpose:** Pregnancy loss ranging from spontaneous abortion (SAB) to stillbirth can result from monogenic causes of Mendelian inheritance. This study evaluated the clinical application of whole exome sequencing (WES) in identifying the genetic etiology for pregnancy loss.

**Methods:** A cohort of 102 specimens from products of conception (POC) with normal karyotype and absence of pathogenic copy number variants were selected for WES. Abnormality detection rate (ADR) and variants of diagnostic value correlated with SAB and stillbirth were evaluated.

**Results:** WES detected six pathogenic variants, 16 likely pathogenic variants, and 17 variants of uncertain significance favor pathogenic (VUSfp) from this cohort. The ADR for pathogenic and likely pathogenic variants was 22% and reached 35% with the inclusion of VUSfp. The ADRs of SAB and stillbirth were 36% and 33%, respectively. Affected genes included those associated with multi-system abnormalities, neurodevelopmental disorders, cardiac anomalies, skeletal dysplasia, metabolic disorders and renal diseases.

**Conclusion:** These results supported the clinical utility of WES for detecting monogenic etiology of pregnancy loss. The identification of disease associated variants provided information for follow-up genetic counseling of recurrence risk and management of subsequent pregnancies. Discovery of novel variants could provide insight for underlying molecular mechanisms causing fetal death.

## INTRODUCTION

The incidence of pregnancy loss from implantation to clinically recognized spontaneous abortions (SAB) was approximately 30%.^1^ From a household survey of women aged 15-44 years in the United States during the 1990-2011, self-reported pregnancy loss was noted in approximately 20% of pregnancies with a trend of an increased risk of 1% per year after excluding maternal factors.^2^ Pregnancy loss includes spontaneous abortion (SAB) or miscarriage defined as fetal death prior to 20 weeks of gestation, and stillbirth defined as fetal death at 20 weeks of gestation or greater. Pregnancy loss can have significant physiologic and psychological consequences for women and families. Identify underlying genetic causes could reduce self-blame among those losing a pregnancy and allow for effective clinical management of future pregnancies.^3^

Genetic abnormalities are known to cause pregnancy loss. Diagnostic karyotyping and microarray analysis identified numerical and structural chromosomal abnormalities and pathogenic copy number variants (pCNVs) in 50% and 4% of products of conception (POC) cases, respectively.^4^ Further analysis of these chromosomal abnormalities and pCNVs identified candidate critical genes and potential interactive gene networks affecting early embryonic development.^4,5^ The first report of whole exome sequencing (WES) on a family with recurrent pregnancy loss due to nonimmune hydrops fetalis identified a homozygous rare variant in a highly conserved region of the CHRNA1 gene as a Mendelian cause.^6^ Further application of WES detected relevant alterations in four out of seven cases of fetal demises,^7^ and compound heterozygous mutations in two out of four miscarriages.^8^ More recent WES studies detected positive, possible and candidate variants in 20%, 45% and 9% of 84 fetal death cases with ultrasound anomalies, respectively,^9^ and sequence variants in 15 out of 19 POC cases with missed abortion.^10^ Targeted sequencing on a panel of 70 genes associated with cardiac channelophies and cardiomyopathies detected pathogenic variants in 12% of 290 cases of stillbirth.^11^ Despite the differences in case selection criteria and inconsistency in variant classification from these studies, accumulated research data indicated that exome sequencing may be instrumental in identifying monogenic causes of a significant portion of pregnancy loss cases and should be integrated into the diagnostic practice.

We performed WES on a cohort of POC samples with normal chromosome and microarray findings. The technical feasibility and sensitivity of this assay were evaluated. Variant classification followed current standards and guidelines developed by the American College of Medical Genetics and Genomics (ACMG) and the Association for Molecular Pathology.^12^ Result interpretation was given by disease association and the possibility of prenatal lethality resulting in pregnancy loss. Approaches to integrate WES into current prenatal diagnosis, genetic counseling and reproduction management were proposed.

## MATERIALS AND METHODS

### Ethics Statement

This study used surplus DNA from POC samples with no personally identifiable private information, it was deemed exempt from ethical oversight and human subject regulations by Yale University Institutional Review Board.

### Selection of POC cases

Specimens of POC were submitted to Yale Clinical Cytogenetics Laboratory. Sequential karyotyping and microarray analyses were performed to detect chromosome abnormalities and pCNVs.^13^ A cohort of 102 cases with normal karyotype and absence of pCNVs collected during 2015 to 2018 were deidentified and selected for WES analysis based on quality and quantity of the leftover DNA. Maternal age, gestational age, clinical indications, gender, and pathologic findings of selected cases were reviewed by a multidisciplinary panel including expertise from Medical Genetics, Obstetrics and Gynecology and Pathology.

### Exome sequencing, variant filtering, classification and confirmation

DNA samples obtained from each case in the cohort were subjected to WES at the Yale Center for Genome Analysis. An initial clinical application of WES estimated the sensitivity for detecting heterozygous variants to be 98% with mean 40X sequencing coverage.^14^ A pipeline to assess the pathogenicity and causality of detected variants was validated as previously described^15^ and adopted for current study. Briefly, paired end sequence reads were converted to FASTQ format and were aligned to the reference human genome assembly GRCh37/hg19 (https://genome.ucsc.edu/). Following variant annotation using a Genome Analysis Toolkit (GATK) and AnnoVar, filtering was applied against allele frequencies and disease citations using databases including Genome Aggregation Database (gnomAD, https://gnomad.broadinstitute.org/), an internal database of Yale DNA Diagnostics Laboratory, ClinVAR (https://www.ncbi.nlm.nih.gov/clinvar/), OMIM (https://www.omim.org) and other in silico attributes. Variants with allele frequency above 3% were excluded from further analysis.

Following ACMG standards and guidelines, the variants with an allele frequency <3% were categorized as pathogenic, likely pathogenic, variant of uncertain significance (VUS), likely benign and benign variants.^12^ For this cohort, an additional criterion of VUS favor pathogenic (VUSfp) was applied by in-house rules to include primarily rare missense variants in a gene with a low rate of benign variants and nearby missense variants reported as disease-causing variants.^16^ Additional deleterious effects of these VUSfp were further analyzed by in-silico tools of PolyPhen, SIFT, and CADD. Pathogenic, likely pathogenic and VUSfp were considered variants of diagnostic value. VUSs and benign or likely benign variants were excluded from further analysis. Variants of diagnostic value were verified by Sanger sequencing. Primers flanking the variants for polymerase chain reaction (PCR) are listed in Supplementary Table S1. Purified PCR products were submitted to the Yale DNA sequencing facility for Sanger sequencing. The scheme of variant annotation, filtering and classification is shown in Figure 1.

**Figure 1.**
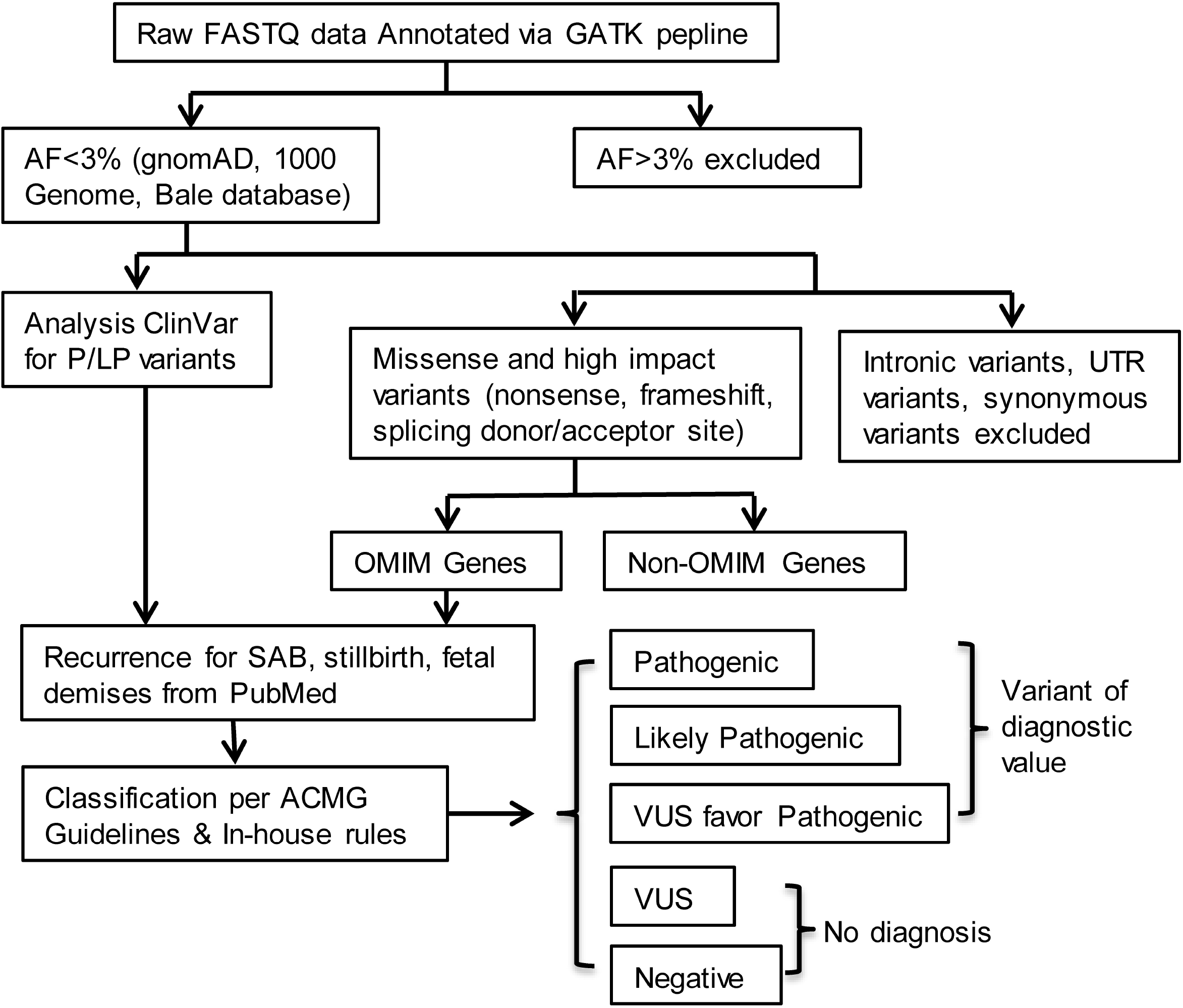
A scheme for variant annotation, filtering and classification. Variants classified as pathogenic, likely pathogenic and VUSfp were considered diagnostic for pregnancy loss.

### Assessment of clinical utility and genetic etiology

Clinical utility of WES for pregnancy loss was assessed by evaluating the abnormality detection rate (ADR) in this cohort. Briefly, the overall ADR was measured as the number of cases with variants of diagnostic value divided by the number of cases analyzed. The ADR in the SAB group (fetal death < 20 weeks of gestation) was compared to the ADR in the stillbirth group (fetal death >= 20 weeks of gestation). Proportions of cases with variants of diagnostic value were assessed in different maternal age groups and trimesters of pregnancy. Pathogenicity and causality of variants of diagnostic value were further examined by their OMIM disease association, inheritance mode and disease categories. Furthermore, a list of reference genes and variants was generated to include 147 genes reported in cases of fetal death throughout gestation from a literature search (Supplementary Table S2).^6-9,11,17-24^ Variants of diagnostic value in this study were compared to the list of reference genes to assess recurrence at either gene level or variant level.

## RESULTS

### Clinical Indications and pathologic findings of the POC cases

Maternal age, gestational age, clinical indications and pathologic findings for these 102 cases were summarized in Supplementary Table S3. The male to female ratio was 49 to 53. The distribution of cases by maternal age groups from age 18-48 years and trimester of pregnancy is shown in Figure 2a and 2b. There were 47 cases of SAB, 45 cases of stillbirth, and 10 cases with unknown gestational age (Figure 2c). Approximately 74% (75/102) of the cases had a maternal age within 26 to 40 years (mean maternal age of 32 years), and 72% (66/92) of the specimens were from the second trimester (mean gestational age of 19 weeks).

**Figure 2.**
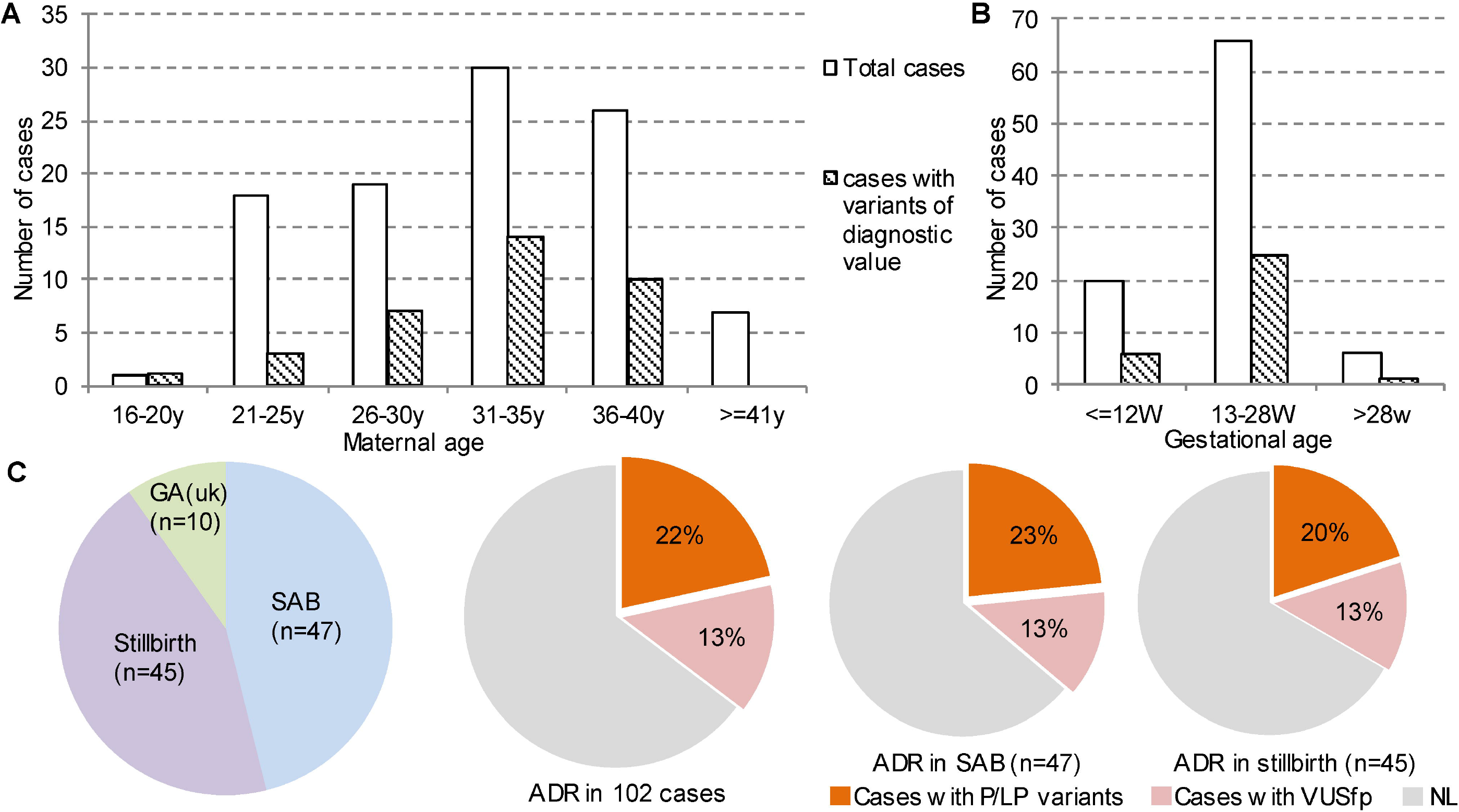
Clinical demographics for the 102 POC cases. A) Distribution of cases by maternal age (open bar) and cases with variants of diagnostic value (shade bar), B) Distribution of case by gestational trimesters (open bar) and cases with variant of diagnostic value (shade bar), C) Number of cases in SAB, stillbirth and unknown gestational ages and ADR for overall cases and for cases of SAB and stillbirth. (uk, unknown; ADR, abnormality detection rate; P/LP pathogenic/likely pathogenic; VUSfp, variants of uncertain significance favor pathogenic, NL normal).

### Variants of potential diagnostic value and abnormality detection rate

Following variant filtering and classification, 39 variants in 36 genes reaching diagnostic value were identified in 36 cases. The six pathogenic variants, 16 likely pathogenic variants, and 17 VUSfp and their OMIM disease association and disease categories are summarized in Tables 1 and 2. Sanger sequencing performed on 26 variants showed 100% consistency with WES sequence results; representative sequencing results of four variants are shown in Supplementary Figure S1. The ADR as measured by the number of cases with pathogenic or likely pathogenic variants divided by the number of cases analyzed was 22% (22/102). With the addition of 14 cases with VUSfp, the overall ADR was 35% (36/102) (Figure 2c). The ADR for SAB was 36% (17/47) and 33% (15/45) for stillbirth, without statistically significant difference (p-value of 0.78). The proportion of cases with variants of diagnostic value by maternal age groups and by trimester of pregnancy is shown in Figures 2a and 2b. The percentage of abnormal cases in maternal ages of 21-25, 26-30, 31-35 and 36-40 years was 17%, 37%, 47% and 38%, respectively. The distribution of paternal ages was not available. The percentage of abnormal cases in first, second and third trimester was 30%, 38% and 17%, respectively. For cases with variants of diagnostic value, 36% (13/36) were detected in pregnant women age 35 years or older and at least 69% (25/36) occurred in the second trimester.

**Table 1.** Pathogenic and likely pathogenic variants detected in this study**\***.

| Case ID | Indications | Gestational age (wks) | Gene | Variant | Zygosity | AF (gnomAD) | Type of Variant | ACMG Classification | OMIM Diseases & Inheritance | Diseases Category |
| --- | --- | --- | --- | --- | --- | --- | --- | --- | --- | --- |
| PL001 | fetal demise | 21 5/7 | NPHS1 | NM_004646:c.2905delC, p.(Leu969Cysfs*32) | Het | 1/30938 | FS | LP | Nephrotic syndrome, type 1; AR | Kidney disease |
|  |  |  |  | NM_004646:c.3110-5C>T | Het | 1/6929 | SpR | VUS |  |  |
| PL003 | fetal anomaly | 22 2/7 | NEFL | NM_006158:c.493G>T, p.(Glu165*) | Het | 0 | Non | LP | Charcot-Marie-Tooth disease, type 1F; AD/AR | Neuropathy |
| PL006 | fetal anomaly of cystic hygroma | 11 | CHD7 | NM_017780:c.5428C>T, p.(Arg1810*) | Het | 0 | Non | LP | CHARGE syndrome; AD | Multi-system |
| PL024 | fetal anomaly | 20 5/7 | GFM1 | NM_024996:c.914delG, p.(Gly305Glufs*6) | Het | 0 | FS | LP | Combined oxidative phosphorylation deficiency 1; AR | Enzyme/Metabolic |
|  |  |  |  | NM_024996:c.1865A>G, p.(Asn622Ser) | Het | 1/141341 | Mis | VUS |  |  |
| PL031 | fetal demise | unknown | DCHS1 | NM_003737:c.6421C>T, p.(Arg2141*) | Het | 0 | Non | LP | Van Maldergem syndrome 1, AR; Mitral Valve Prolapse 2; AD | Cardiac anomaly |
| PL033 | fetal demise | unknown | PKD1 | NM_001009944:c.12124C>T, p.(Gln4042*) | Het | 0 | Non | P | Polycystic kidney disease 1; AD | Kidney disease |
| PL043 | missed abortion | 8 3/7 | AUTS2 | NM_015570:c.53C>A, p.(Ser18*) | Het | 0 | Non | LP | Mental retardation, autosomal dominant 26; AD | Neurodevelopmental disorder |
| PL048 | fetal demise | 15 | GATA4 | NM_002052:c.1325C>T, p.(Ala442Val) | Het | 1/62871 | Mis | LP | GATA4-associated heart disease (OMIM:600576), AD | Cardiac anomaly |
| PL052 | missed abortion | 10 4/7 | FGFR3 | NM_000142:c.1537G>T, p.(Asp513Tyr) | Het | 0 | Mis | LP | LADD syndrome; AD | Multi-system |
| PL055 | fetal anomaly | 22 4/7 | PIK3R2 | NM_005027:c.1117G>A, p.(Gly373Arg) | Het | 0 | Mis | P | Megalencephaly-polymicrogyria-polydactyly-hydrocephalus syndrome 1; AD | CNS abnormality |
| PL057 | fetal anomaly | 13 3/7 | SMPD1 | NM_000543:c.1133G>A, p.(Arg378His) | Homo | 1/14029 | Mis | P | Niemann-Pick disease, type A/B ; AR | Enzyme/meta bolic |
| PL058 | fetal anomaly | 19 4/7 | GPD1L | NM_015141:c.372A>G, p.(Ile124Met) | Het | 1/20196 | Mis | LP | Brugada syndrome 2; AD | Arrhythmia |
| PL059 | fetal demise | 26 | F11 | NM_000128.3:c.1716+1G>A | Het | 1/125746 | SpD | P | Factpr XI deficiency; AD/AR | Coagulation |
| PL060 | Spina bifida | 19 | FOXP2 | NM_014491.3:c.258+1G>A | Het | 1/250396 | SpD | LP | Speech-language disorder-1; AD | Neurodevelop mental disorder |
| PL061 | fetal demise | 32 | SETD5 | NM_001080517:c.4106dupT, p.(Ser1370Glufs*10) | Het | 1/124629 | FS | LP | Mental retardation, autosomal dominant 23; AD | Neurodevelop mental disorder |
| PL066 | fetal demise | 26 | ATP7B | NM_000053:c.3207C>A, p.(His1069Gln) | Homo | 1/1981 | Mis | P | Wilson Disease; AR | Enzyme/Meta bolic |
| PL067 | Skeletal dysplasia | 20 | FGFR3 | NM_000142:c.742C>T, p.(Arg248Cys) | Het | 0 | Mis | P | Thanatophoric dysplasia, type I ; AD | Skeletal dysplasia |
| PL072 | fetal demise | 22 | SCN5A | NM_198056:c.5393G>A, p.(Trp1798*) | Het | 0 | Non | LP | SCN5A-associated cardiac diseases, AD | Arrhythmia |
| PL073 | fetal anomaly | 18 6/7 | GREB1 L | NM_001142966:c.1305dupA, p.(Asp436Argfs*32) | Het | 0 | FS | LP | Renal hypodysplasia/aplasia 3; AD | Kidney disease |
| PL076 | embryonic demise | 7 | SMAD6 | NM_005585:c.652C>T, p.(Gln218*) | Het | 0 | Non | LP | Aortic Valve Disease, AD | Cardiac anomaly |
| PL078 | fetal demise | 16 | KIAA1109 | NM_015312:c.3986A>G, p.(Tyr1329Cys) | Het | 1/31209 | Mis | LP | Alkuraya-Kucinkas syndrome; AR | Multi-system |
|  |  |  |  | NM_015312:c.11093C>G, p.(Ser3698Cys) | Het | 1/69995 | Mis | VUS |  |  |
| PL088 | fetal anomaly | 18 | NEB | NM_001271208:c.24094C>T, p.(Arg8032*) | Het | 1/2052 | Non | LP | Nemaline myopathy 2, AR | Myopathy |
|  |  |  |  | NM_001271208:c.20098C>A, p.(Leu6700Ile) | Het | 1/2336 | Mis | VUS |  |  |

\* AF: Allele Frequency, Het: Heterozygous, Homo: Homozygous, Non: Nonsense, Mis: Missense, FS: Frameshift, SpR: Splicing receptor site, SpD: Splicing donor site, P: Pathogenic, LP: Likely pathogenic, AD: autosomal dominant inheritance, AR: autosomal recessive inheritance.

**Table 2.** Variants of uncertain significance favor pathogenic detected in this study**\***.

| Case ID | Indications | Gestational age (wks) | Gene | Variant | Zygosity | AF (gnomAD) | Type of Variant | OMIM Diseases & Inheritance | Diseases Category |
| --- | --- | --- | --- | --- | --- | --- | --- | --- | --- |
| PL005 | fetal demise | 13 | FGFR2 | NM_000141:c.182G>A, p.(Arg61His) | Het | 1/61546 | Mis | FGFR2-related syndrome , AD | Skeletal dysplasia |
| PL019 | Fetal demise, multiple anomalies | 22 4/7 | FBN1 | NM_000138:c.7999G>A, p.(Glu2667Lys) | Het | 1/30250 | Mis | Marfan syndrome ; AD | Multi-system |
| PL029 | embryonic demise | 10 | MIB1 | NM_020774:c.351dupA, p.(His118Thrfs*16) | Het | 0 | FS | Left ventricular noncompaction 7; AD | Cardiac anomaly |
|  |  |  | SHOC2 | NM_007373:c.241A>G, p.(Thr81Ala) | Het | 1/35826 | Mis | Noonan syndrome-like with loose anagen hair; AD | Rasopathy |
| PL032 | fetal demise | 22 | HACE1 | NM_020771:c.212T>C, p.(Ile71Thr) | Homo | 0 | Mis | Spastic paraplegia and psychomotor retardation with or without seizures; AR | Neurodevelopmental disorder |
| PL034 | missed abortion | 18 | NIPBL | NM_133433:c.1927A>G, p.(Lys643Glu) | Het | 0 | Mis | Cornelia de Lange syndrome 1; AD | Multi-system |
| PL037 | fetal demise | unknown | SOS1 | NM_005633:c.205G>A, p.(Asp69Asn) | Het | 0 | Mis | Noonan Syndrome 4; AD | Rasopathy |
|  |  |  | TTC21B | NM_024753:c.511G>A, p.(Gly171Arg) | Homo | 1/3400 | Mis | Short-rib thoracic dysplasia 4 with or without polydactyly; AR | Multi-system |
| PL038 | missed abortion, fetal anomalies | 23 6/7 | TSC2 | NM_000548.3:c.1946+5G>T | Het | 0 | SpR | Tuberous sclerosis-2; AD | Multi-system |
| PL039 | fetal demise | 21 2/7 | SYNGAP1 | NM_006772:c.773G>A, p.(Arg258His) | Het | 1/81822 | Mis | Mental retardation, autosomal dominant 5; AD | Neurodevelopmental disorder |
| PL071 | fetal anomaly | 21 | COQ2 | NM_015697:c.662T>C, p.(Leu221Pro) | Homo | 1/124388 | Mis | Coenzyme Q10 deficiency, primary, 1 ; AR | Enzyme/Metabolic |
| PL075 | PRROM | 20 5/7 | SCN5A | NM_198056:c.1933T>G,<br>p.(Ser645Ala) | Het | 0 | Mis | SCN5A related<br>disorders; AD/AR | Arrhythmia |
| PL080 | fetal demise | unknown | JAG1 | NM_000214:c.218C>G,<br>p.(Thr73Arg) | Het | 0 | Mis | Alagille syndrome 1,<br>AD; Tetralogy of<br>Fallot, AD | Multi-system |
| PL083 | fetal demise | 14 3/7 | IDS | NM_000202:c.778C>T,<br>p.(Pro260Ser) | Hemi | 0 | Mis | Mucopolysaccharidos<br>is II; XLR | Enzyme/Met<br>abolic |
| PL090 | fetal demise | 11 3/7 | CSRP3 | NM_003476:c.3G>A, p.? | Het | 0 | SL | Cardiomyopathy,<br>dilated, 1M ; AD | Cardiac<br>anomaly |
|  |  |  | NIPBL | NM_133433:c.193C>G,<br>p.(Leu65Val) | Het | 0 | Mis | Cornelia de Lange<br>syndrome 1; AD | Multi-system |
| PL094 | fetal demise | 15 1/7 | COL1A1 | NM_000088:c.4024G>A,<br>p.(Gly1342Ser) | Het | 1/81771 | Mis | Osteogenesis<br>imperfecta I; AD | Skeletal<br>dysplasia |

\* AF: Allele Frequency, Het: Heterozygous, Homo: Homozygous, Hemi: Hemizygous, Mis: Missense, FS: Frameshift, SpR: Splicing receptor, SL: StartLoss, AD: autosomal dominant inheritance, AR: autosomal recessive inheritance.

### Inheritance mode and disease categories

Among the 39 variants of diagnostic value, 67% (26/39) were associated with an autosomal dominant (AD) condition, 21% (8/39) with an autosomal recessive (AR) condition, 10% (4/39) with a condition that can be either AD or AR, and 3% (1/39) with an X-linked recessive condition. Four VUSs were also listed in Table 1 as each one of them was found together with one pathogenic or likely pathogenic variant in the same gene. However, the phasing of the VUS and the variant of diagnostic value in the AR gene was undetermined without follow up parental studies. Three cases had variants in more than one gene implicated. Variants associated with dominant conditions had zero to extremely low allele frequency in the general population, and were absent in patients tested for other indications in the Yale DNA Diagnostics Laboratory.

Twelve disease categories were noted for the 36 genes carrying variants of diagnostic value. The most prevalent disease category was multi-system disorders, which included eight genes *CHD7, FGFR3, JAG1, TSC2, KIAA1109, TTC21B, FBN1* and *NIPBL* (2) in nine cases. Additionally, variants in genes *SOS1* and *SHOC2* of RASopathy, which is a specific group of multi-system disorders, were identified in two cases. The second most common category was cardiac anomalies involving five genes *GATA4, DCHS1, SMAD6, MIB1* and *CSRP3* in five cases, and arrhythmia involving two genes *GPD1L* and *SCN5A* (2) in three cases. Variants in five genes *AUST2, FOXP2, SETD5, HACE1* and *SYNGAP1* associated with neurodevelopmental disorders were identified in five cases. Variants in five genes *ATP7B, COQ2, IDS, SMPD1* and *GFM1* for enzyme and metabolic diseases were noted in five cases. Three cases had variants in genes *NPHS1, PKD1* and *GREB1L* associated with kidney diseases. Variants in genes *FGFR2, FGFR3* and *COL1A1* associated with skeletal dysplasia were identified in three cases. Variants were also identified in other disease categories including neuropathy (*NEFL*), myopathy (*NEB*), coagulation (*F11*) and CNS abnormality (*PIK3R2*) in four cases.

Fifteen genes of 17 cases from this cohort, including *CHD7, COL1A1, CSRP3, FBN1, FGFR2, FGFR3, GATA4, GPD1L, GREB1L, NIPBL, NPHS1, PIK3R2, SCN5A, SOS1* and *TSC2* had been previously reported in separate case series in association with fetal death as listed in Supplementary Table S2. Among these genes, variants in the *FGFR3, SCN5A* and *NIPBL* genes were identified in two cases each within this cohort. This result indicated that 42% (15/36) of the genes involving 47% (17/36) of cases showed recurring association with fetal death. Recurrence at variant level was also observed in three cases. Variant p.(Gly373Arg) in the *PIK3R2* gene of case PL055 with fetal arthrogryposis was previously reported in a fetus with bilateral ventriculomegaly and lissencephaly in a trio-exome study.^23^ Variant p.(Ile124Met) in the *GPD1L* gene was seen in a SAB case PL058 with single left kidney and two vessel cords; a variant at the same amino acid p.(Ile124Val) was reported in four cases of still birth.^11^ A known pathogenic variant for Thanatophoric dysplasia, p.(Arg248Cys) in the *FGFR3* gene identified in case PL067, was reported previously in a case of fetal demise.^24^

## DISCUSSION

Clinical WES was first introduced in 2009 and was adopted quickly as a highly effective approach for postnatal and prenatal genetic diagnosis of Mendelian disorders.^14,15,22^ A retrospective analysis of WES on 146 fetuses with ultrasound abnormalities demonstrated a diagnostic rate of 32% for Mendelian autosomal dominant and recessive disorders.^22^ A prospective prenatal exome sequencing analysis of a panel of 1628 genes for developmental disorders on a cohort of 610 fetuses with ultrasound detected structural anomalies showed a diagnostic rate of 12% for variants of pathogenicity and potential clinical usefulness.^24^ The ADR of this cohort was 22% for pathogenic and likely pathogenic variants, with an additional 14% for VUSfp. This diagnostic yield was comparable to the 12%-32% from prenatal clinical exome sequencing and the 20%-74% from deceased fetuses with ultrasound anomalies.^9^ The ADR of 36% from SAB was slightly more than that of 33% from stillbirth, but the difference was not significant. Combing findings from karyotyping, microarray analysis and WES, the ADR from POC specimens was 50%, 4% and 22%-36% for chromosomal abnormalities, pCNVs and monogenic variants, respectively.^4^ These results indicated that, with the addition of WES, current genetic testing can identify a specific genetic etiology in about three quarters of the pregnancy loss cases. Since WES can also detect aneuploidies and pCNVs except for rare balanced or derived chromosome rearrangements and low-level mosaicisms, WES could replace routine cytogenetic evaluations with karyotype and microarray to detect most genetic abnormalities in pregnancy loss.

The challenge in the interpretation of monogenic genetic contributions to pregnancy loss is to clarify disease association and establish cause-effect relationship for fetal death. Recently, accumulated evidence supported causative genetic variants recurring in essential genes involved in embryonic development, organ development and various functions.^25-28^ All the variants of diagnostic value reported in this cohort have a clear association with an OMIM condition. Dominant lethal conditions are poorly understood in humans. In this study, approximately 67% of the variants of diagnostic value were identified in an AD condition. Actually, two AD conditions were noted in case PL029 with one VUSfp each in the *MIB1* and *SHOC2* genes and case PL090 with one VUSfp each in the *CSRP3* and *NIPBL* genes. Similarly, case PL037 had one homozygous VUSfp in the *TTC21B* which is a recessive ciliopathy gene and a VUSfp variant in the *SOS1* gene for a dominant RASopathy. It is possible that the variants in both genes act synergistically to result in fetal death. Of the eight cases associated with AR conditions, homozygous pathogenic variants or VUSfps in the *SMPD1, ATP7B, HACE1, TTC21B* and *COQ2* genes were detected in five cases. While adverse effect on fetal development is more apparent in four genes, *ATP7B* associated Wilson disease as the cause of fetal death will require further evidence. These cases suggested a possible founder effect in the local testing population. Heterozygous variants of likely pathogenic and VUS were detected in the *NPHS1, KIAA1109* and *NEB* genes associated with a AR condition in three cases. Follow up parental study to determine the phase of these variants and functional analysis on gene expression will be necessary to assess their association to fetal death. A very rare VUSfp in the *IDS* gene associated with X-linked Mucopolysaccharidosis II was identified in a male SAB case; its pathogenicity of glycosaminoglycan accumulation in CNS and other organ systems for fetal death would need further investigation on fetal autopsy. Some of the variants reported are in genes associated with adult onset diseases, such as *FBN1, PKD1* and *SMAD6*. A variant in the *FBN1* gene and a 41-kb deletion containing the *TSC2* and *PKD1* genes have been reported in fetuses with ultrasound detected abnormality.^22,24^ These findings raise the possibility that adult onset conditions associated with these genes may have a broader phenotype than we recognized, and could affect fetal development. Likewise, identifying pathogenic variants in genes known to be associated with neurodevelopmental disorders, such as the *AUTS2, FOXP2, SETD5* and *SYNGAP1* genes, raises a similar possibility of a broader impact of these genes on fetal development. The results from this cohort illustrated that detailed clinicopathologic investigation is needed to determine the cause of fetal death.

Recurrence of certain disease categories and related genes and variants are considered clinical evidence supporting causal effect for pregnancy loss. In this cohort, disease categories showed considerable overlap with those recognized in prior studies including multisystem disorders, cardiovascular abnormalities, urinary system abnormalities, skeletal dysplasia, and central nervous system abnormalities.^9,11,22-24^ Approximately 47% of the cases in this cohort had evidence supporting the identification of a genetic cause for fetal death from previous studies. Of the 15 recurrent genes, most are associated with multisystem disorders, including the *CHD7, FBN1, FGFR3, NIPBL* and *SOS1* genes; and the second most common ones are associated with cardiac anomalies or arrhythmia including the *CSRP3, GATA4, GPD1L* and *SCN5A* genes. The other genes are associated with skeletal dysplasia *(COL1A1, FGFR2* and *FGFR3)*, kidney diseases (*GREB1L* and *NPHS1)* and CNS abnormality *(PIK3R2*). More specifically, the variant identified in the *PIK3R2* gene, p.(Gly373Arg), is identical to the previously reported *PIK3R2* variant,^23^ and a variant causing Thanatophoric dysplasia, p.(Arg248Cys) in the *FGFR3* gene, was previously reported in a case of fetal demise.^24^ Variants affecting the same amino acid Ile124 were identified in the *GPD1L* gene.^11^ Such recurrence at the variant level further supports the important function of certain amino acid positions and protein structure, and that genetic abnormalities at these positions likely contributed to fetal death. The fact that there are recurrent disease categories, genes, and variants found in different studies supports the presence of shared mechanisms and monogenic contributions to pregnancy loss. These results also highlighted that although the genetic etiology underlying pregnancy loss appears to be diverse and heterogenous, some genes may play more critical roles for normal fetal development than others, and fetal development cannot be sustained once these essential genes are deregulated.

There were a few limitations in this study and thus approaches overcoming these limitations should be implemented for clinical use and further research. First, this cohort of 102 cases was adequate to validate the clinical utility of WES but the sample size was insufficient to have a comprehensive evaluation of genetic etiology for pregnancy loss. Given the heterogeneity of disease categories presented in this cohort and other studies, collaborative studies on a large case series will be needed to investigate the monogenic etiology of fetal death. Second, the samples used for this study were de-identified and follow-up parental study was not pursued. Reanalysis of phase and parental origin of variants could be performed following current ACMG recommendations.^29^ A more complete fetal clinical evaluation combined with a trio study design or a study with follow up parental testing would allow for better assessment of the causality of the variants. Trio analysis has been shown to improve diagnostic yield and variant interpretation over singleton study.^9^ Finally, this study only focused on OMIM genes and was not designed to evaluate non-OMIM genes that might be important for fetal growth and development. Trio study will be critical for discovery of new genes important to fetal development. Furthermore, studies using cellular and animal models should be an integral component of future study design to clarify the functional impact of the identified variants, especially the recurrent and novel variants, on fetal death.

In conclusion, this study demonstrated that 22%-35% of pregnancy losses have variants of diagnostic value in genes that may contribute to fetal death, supporting the use of WES as a valuable genetic testing tool in identifying a cause for pregnancy loss. Identification of recurrent genes and variants associated with fetal death may lead to better understanding of the functions of known OMIM genes in fetal development and their roles in pregnancy loss. Identification of variants of diagnostic value provides necessary information for recurrence risk assessment, follow up parental studies, prenatal genetic counseling, and management of subsequent pregnancies.

## Data Availability

The online version of this article contains supplementary figure S1 and supplementary tables S1, S2 and S3.

